# Time at the curb, not the rate of head scanning, distinguishes street crossing in virtual reality with severe-to-profound visual impairment

**DOI:** 10.64898/2026.11.04.26367282

**Authors:** Batuhan Erkat-Cohen, Dinesh Venugopal, Roksana Sadeghi, Chau Tran, Will Gee, Brittnee Livingston, Gislin Dagnelie, Arathy Kartha

**Affiliations:** SUNY College of Optometry, New York, New York, United States; Wilmer Eye Institute, Johns Hopkins University School of Medicine, Baltimore, Maryland, United States; Department of Biomedical Engineering, Johns Hopkins University, Baltimore, Maryland, United States; BaltiVirtual, Baltimore, Maryland, United States; Central Association for the Blind and Visually Impaired, Utica, New York, United States

**Keywords:** severe-to-profound visual impairment, street crossing, head movements, virtual reality, orientation and mobility, reaction time

## Abstract

**Purpose:** We tested whether people with severe-to-profound visual impairment compensate at street crossings with more head scanning and/or more time at the curb.

**Methods:** Ten normally sighted controls, tested with normal vision and with Bangerter filters simulating severe-to-profound visual impairment (approximately 20/2000), and 17 adults with severe-to-profound visual impairment crossed a virtual street in a head-mounted display. Four complexity levels ranged from an empty street to yielding vehicles with pedestrian avatars; only the two most complex levels contained moving vehicles. Cumulative absolute head yaw and latency to step-off were analyzed with mixed-effects models, and latencies were fitted with an ex-Gaussian distribution.

**Results:** Compared with normal vision, latency to step-off was 1.57 times longer with simulated and 1.82 times longer with clinical severe-to-profound visual impairment (Bonferroni *p* < 0.001 and *p* = 0.008, respectively). In severe-to-profound visual impairment, the additional latency was attributable to the slow tail of the latency distribution (exponential component τ longer by 3.40 s without and 6.20 s with moving vehicles; Bonferroni *p* = 0.037 and 0.022, respectively); peak position did not differ significantly. Before step-off, cumulative absolute head yaw increased with complexity, *F*_(3, 115.8)_ = 37.16, *p* < 0.001, and was 1.88 times higher with simulated impairment than with normal vision (Bonferroni *p* = 0.024); it did not differ significantly between clinical impairment and normal vision, either before step-off or during walking (both Bonferroni *p* ≥ 0.479). During walking, its increase with complexity depended on viewing condition, *F*_(6, 109.3)_ = 5.23, *p* < 0.001.

**Conclusions:** Severe-to-profound visual impairment was associated with longer and more variable time at the curb, without a significant increase in cumulative head scanning, although the increase in head scanning with complexity during walking was attenuated. Time at the curb is relevant to orientation and mobility practice and accessible crossing design.

**Significance:** Pedestrian injuries are most severe at uncontrolled crossings. At a virtual crossing without tactile and auditory cues, adults with severe-to-profound visual impairment took longer than sighted adults to step off the curb. Time spent at the curb may inform orientation and mobility assessment and accessible crossing design.

## INTRODUCTION

Pedestrians account for an increasing proportion of traffic fatalities in the United States ^1^. Pedestrians struck at non-signalized crossings sustain more severe injuries and have higher mortality than those struck at signalized crossings ^2^. At an uncontrolled crossing, no signal is available, and the decision to initiate crossing depends entirely on the pedestrian’s own assessment of the traffic.

Severe vision loss is associated with a greater risk of death from unintentional injury ^3^. For example, pedestrians with severe peripheral field loss identify fewer crossable gaps and more often accept gaps that are too short ^4^. However, pedestrians with low vision who travel independently judge crossing gaps as accurately as normally sighted pedestrians; without visual cues, both groups judge them less accurately ^5^. These findings can be reconciled if vision loss limits the acquisition of gap information rather than its evaluation. This question remains unresolved in people with severe-to-profound visual impairment (SPVI), whose residual vision is limited and, in ultra-low vision, rudimentary at best ^6^. Yet in this population, mobility is among the activities for which people report benefiting from their residual vision ^7^. The open question is therefore not whether crossing outcomes differ with vision loss, but how the information required for crossing is acquired.

Orientation and mobility training teaches pedestrians with low vision to look in both directions before entering the roadway. If degraded vision provides less information per look, visually impaired pedestrians would be expected to scan more. The clearest evidence of compensatory scanning comes from homonymous hemianopia, in which pedestrians scan farther and more often toward the blind than toward the seeing hemifield ^8^; in a driving simulator, early large head scans toward the blind hemifield were associated with earlier fixation of, and faster responses to, hazards on that side ^9^. Hemianopia, however, is a special case, because visual acuity is typically good and there is a defined direction toward which to compensate. Without such a direction, the prediction is not supported: pedestrians with age-related macular degeneration, for instance, turn their heads less often than normally sighted pedestrians at a real intersection ^10^. In laboratory gaze tasks, people with early to moderate retinitis pigmentosa increased head movement at the expense of eye movement, without increasing total gaze movement ^11^. Even in hemianopia, the evidence is inconsistent: a virtual street-crossing study comparing clinical, simulated and absent hemianopia found no group differences in scanning behavior ^12^. Importantly, none of these studies examined street crossing at the severe end of visual impairment; in the one that included late-stage retinitis pigmentosa, these participants made minimal gaze movements ^11^. Head and eye movements have been recorded in a head-mounted display while patients with central, peripheral or combined impairment detected approaching vehicles and identified safe crossing opportunities in a virtual street ^13^. In the combined-loss subgroup of that study, the group closest to SPVI, head turns were less frequent than in controls. Because those participants judged a recorded scene while stationary, continuous head-yaw kinematics have not been measured while people with clinical SPVI physically cross a street.

Crossing is a decision made under uncertainty: the pedestrian accumulates information about approaching vehicles and initiates crossing when it is judged safe. Models of decision making predict that the duration of this decision depends both on the rate at which visual information about approaching vehicles is acquired and on the amount of that information required before committing ^14–16^. The time spent at the curb, and its distribution across crossings, may therefore indicate the consequences of degraded vision.

To test this, we compared normally sighted adults, the same adults under simulated SPVI, and adults with clinical SPVI as they crossed a virtual street at graded levels of environmental complexity. The simulation separated the immediate effect of acuity loss from adaptation, orientation and mobility training, and the habits that accompany long-standing vision loss. Because participants completed every crossing, measures of head scanning and of the time spent at the curb characterize how a crossing was approached, not whether the decision was safe. We asked whether people with severe vision loss compensate by increasing head scanning or by spending more time at the curb.

## MATERIALS & METHODS

### Participants

Twenty-seven adults participated: 10 normally sighted controls and 17 participants with SPVI (**Table 1**), all recruited from the Johns Hopkins Department of Ophthalmology. SPVI was defined as visual acuity of 20/200 (1.0 logMAR) or worse, spanning the severe (20/200–20/400) and profound (worse than 20/400, including near-total) categories of the International Classification of Diseases, Ninth Revision, Clinical Modification; the profound category corresponds broadly to ultra-low vision ^7^, for which orientation and mobility assessments have been developed for vision-restoration trials ^17^. Acuity was estimated with the Berkeley Rudimentary Vision Test ^18^. Controls had no ocular disease or surgery and an acuity of 20/25 (0.1 logMAR) or better in each eye. We excluded participants with self-reported cognitive, hearing, or mobility impairment, and participants wore their habitual correction.

**Table 1.** Participants with SPVI: gender, cause of vision loss, age and visual acuity.

| Participant | Gender | Cause of vision loss | Age, y | Visual acuity, logMAR |
|---|---|---|---|---|
| 1 | F | Cone–rod dystrophy | 41–45 | 3.5 |
| 2 | — | — | — | — |
| 3 | M | Diabetic retinopathy and glaucoma | 31–35 | 1.4 |
| 4 | F | Retinopathy of prematurity | 31–35 | 1.2 |
| 5 | F | Marfan syndrome with retinal detachment | 31–35 | 1.4 |
| 6 | M | Retinal detachment secondary to pathologic myopia | 31–35 | 1.4 |
| 7 | M | Glaucoma | 86–90 | 2.0 |
| 8 | M | Retinitis pigmentosa | 46–50 | 1.6 |
| 9 | F | Leber congenital amaurosis | 21–25 | — |
| 10 | M | Retinitis pigmentosa | 36–40 | 1.4 |
| 11 | M | Glaucoma | 71–75 | 1.3 |
| 12 | F | Optic nerve atrophy | 46–50 | 1.3 |
| 13 | M | Retinitis pigmentosa | 56–60 | 3.5 |
| 14 | F | Congenital cataracts and glaucoma | 76–80 | 1.6 |
| 15 | F | Pseudotumor cerebri | 31–35 | 1.8 |
| 16 | F | Cone–rod dystrophy | — | 1.0 |
| 17 | M | Retinitis pigmentosa | 56–60 | — |

Controls were tested under normal vision (NV) and under simulated SPVI (sSPVI), produced with stacked Bangerter filters that reduced acuity to approximately 20/2000 (2.0 logMAR), as in previous work from our laboratory ^19^. The two conditions were run as separate sessions in counterbalanced order (all controls completed both; one session in the NV condition did not have head movement data). Mean age was 47.5 years (*SD* = 18.8; age bands from 21–25 to 86–90 years; 2 not recorded) for SPVI participants, and 8 of the 16 with recorded gender were women. Controls had a mean age of 35.1 years (*SD* = 10.5), and 5 of the 10 were women ^20^. Participants with SPVI completed the protocol with and without a long cane; only trials without a mobility aid (cane or guide dog) were analyzed.

### Apparatus

The virtual environment was developed in Unity and presented on an HTC VIVE Pro Eye head-mounted display (HTC Corporation, New Taipei City, Taiwan; 1440 × 1600 pixels per eye, 110° diagonal field of view, 90 Hz), with head pose tracked in six degrees of freedom by external base stations. Participants physically walked each crossing in the tracked laboratory space (**Figure 1**). An experimenter walked beside the participant in every trial. No auditory or tactile traffic information was provided; the only non-visual cue was a pair of wooden platforms on the floor at the positions of the two virtual curbs.

**Figure 1.**
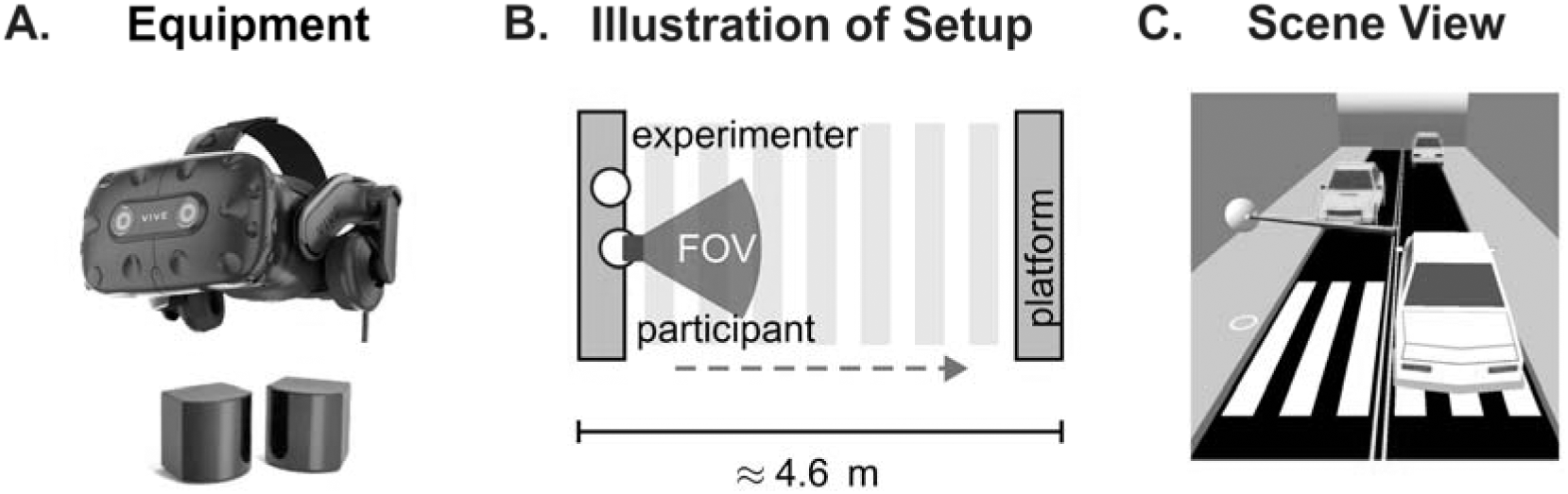
Experimental apparatus, physical testing space, and virtual street environment. **A.** The HTC VIVE Pro Eye head-mounted display used to present the virtual environment. **B.** Plan view of the physical testing space. The dashed arrow indicates the walking path, the shaded wedge the head direction, and the pale floor stripes the virtual crosswalk superimposed on the plan. Wooden platforms at the two virtual curbs were the only tactile cue, and an experimenter walked beside the participant in every trial. The dimension indicates the distance of approximately 4.6 m from the start position to the target on the opposite curb. **C.** The virtual street, rendered from above and behind the start position, which is marked by the sphere at the near curb.

### Virtual environment and task

The environment consisted of a straight roadway with a curb on each side and a marked crosswalk (**Figure 2**). Each trial began with the participant standing on the near curb, facing the crosswalk. Participants could view the scene for as long as they wished before crossing to a target on the opposite curb, approximately 4.6 m away. At levels A and B, the street was one-way without a center line, and at levels C and D, it was two-way with a double yellow center line. Level A was empty; level B contained parked cars and moving pedestrian avatars; level C contained moving vehicles in both directions that yielded at the crosswalk; and level D was level C with moving pedestrian avatars added. For analyses of pairs of levels, levels A and B, which contained no moving vehicles, were grouped as low demand, and levels C and D, which contained moving vehicles, as high demand.

**Figure 2.**
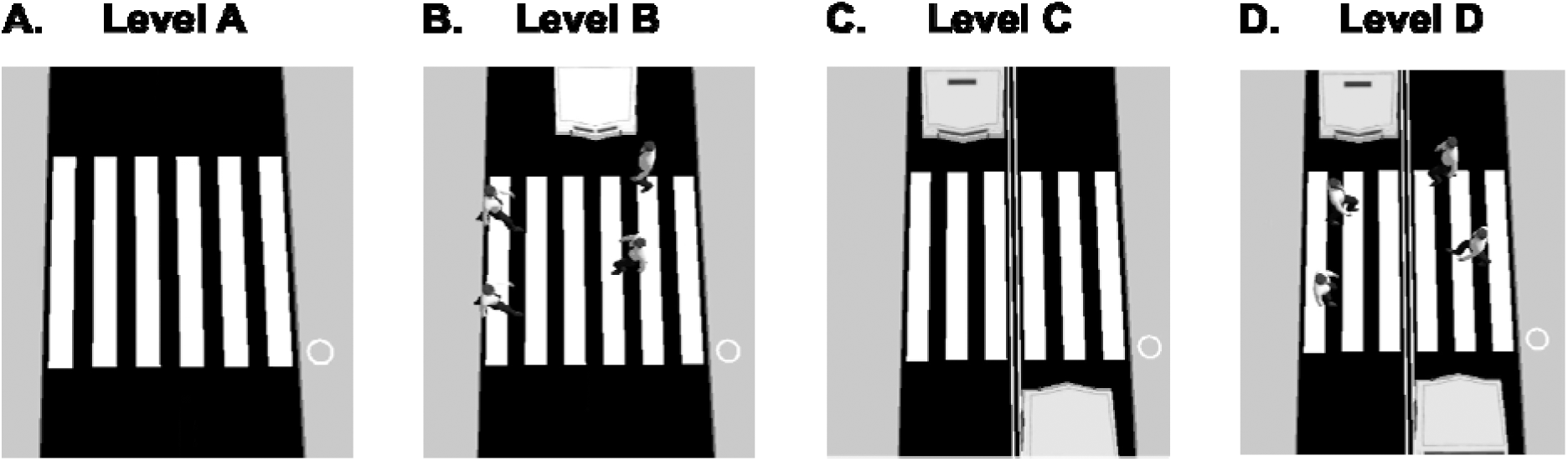
Overhead views of the four graded levels of environmental complexity. Each panel shows one complexity level from directly above, with the roadway running vertically, the crosswalk marked by white stripes, and the participant’s start position at the curb marked by the open circle at the right of each panel. The crosswalk is identical in all four panels. **A.** Level A, the empty one-way street. **B.** Level B, the one-way street with parked cars and moving pedestrian avatars on and beside the crosswalk. **C.** Level C, the two-way street, with a double yellow center line and moving vehicles traveling in both directions, which yielded at the crosswalk. **D.** Level D, the two-way street of level C with moving pedestrian avatars added. Levels were presented in order of increasing complexity (A to D), each preceded by a practice trial. No error representation is shown, because the panels are renderings of the environment, not data.

### Procedure

Street crossing was the first of three virtual environments in each session. Levels were presented in a fixed order from A to D, so that every participant encountered the least demanding environment first. Each level was preceded by an unrecorded practice trial. Participants were instructed to cross at a comfortable speed and to avoid cars and pedestrians. Each session contributed one crossing per level, for a total of 144 trials (NV 36, sSPVI 40, SPVI 68).

### Head-movement recording and preprocessing

Head position (X along the crossing, Y vertical, Z lateral) and rotation were logged at approximately 9.1 Hz. Of the three rotation angles, only yaw, the rotation about the vertical axis, was analyzed. Yaw was referenced to 0° across the crosswalk, with positive values leftward, within −180° to 180°, and was not filtered; sessions that initialized facing the opposite direction (circular mean heading over the first 30 samples outside ±90°) were rotated by 180°. Walking speed was computed as the horizontal (X–Z) displacement of the head between samples divided by the sampling interval, then smoothed with a 10-sample (approximately 1.1 s) moving average.

#### Data cleaning

A guide-dog session for one participant with SPVI was excluded. Where a level was recorded twice, the first recording that reached the far curb, at least 4.3 m from the start, was analyzed; the earlier recordings were a false start and an abandoned attempt. Trials in which the participant reached the far curb and then moved back toward the start by more than 1 m were ended at the farthest point (three trials for one control participant).

### Study Variables

#### Step-off detection and epoch boundaries

Step-off was the first sample at which smoothed walking speed reached 0.3 m/s. The pre-initiation epoch extended from trial onset to the preceding sample, and its duration was the latency to step-off. Latency to step-off agreed with the latency detected by the curb step-down criterion of our recent work ^20^ (a head drop of at least 0.1 m within 0.5 m of the start) in the 136 trials in which a step-down was detected, with high correlation (Spearman *r*_(134)_ = 0.87, Pearson *r*_(134)_ = 0.98, both *p* < 0.001). The two latency distributions did not differ (**Figure S1**). The speed criterion was retained because it marks the onset of sustained walking rather than the step-down. The walking bout was the first run of at least 25 consecutive samples (approximately 2.7 s) above 0.3 m/s. We identified it in 139 of 144 trials (NV 36, sSPVI 40, SPVI 63), and reported its duration as walking-bout duration.

#### Cumulative absolute yaw and scanning rate

The primary measure, cumulative absolute yaw (Ψ_scan_ = Σ|Δψ□|), was the sum of absolute sample-to-sample yaw changes, each wrapped to ±180°, computed over the whole trial, the pre-initiation epoch and the walking bout (**Figure 3**). Cumulative absolute yaw quantifies the total angular excursion of the head irrespective of direction; a 40° turn to the left and then back, for example, contributes 80°. Scanning rate (°/s) was Ψ_scan_ divided by the duration of the corresponding epoch (latency to step-off or walking-bout duration).

**Figure 3.**
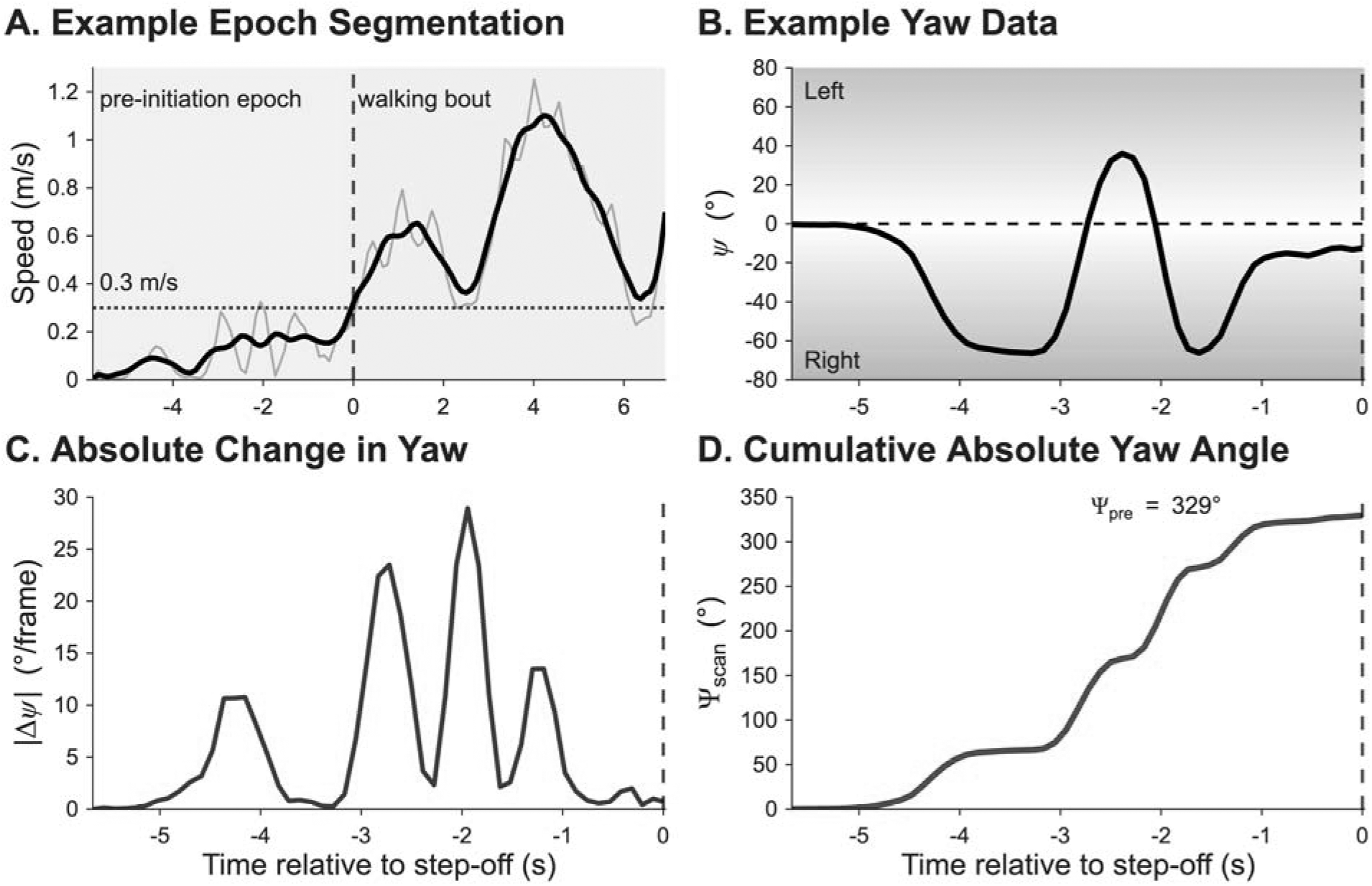
Epoch segmentation and the derivation of cumulative absolute head yaw, illustrated on a single trial. All four panels show the same trial and share a common time origin at step-off (dashed vertical line at 0 s). **A.** Walking speed; the gray trace is the raw per-sample speed and the black trace the same series after a 10-sample moving average. The dotted horizontal line is the 0.3 m/s threshold. The interval before step-off, shaded blue, is the pre-initiation epoch; the first contiguous run of at least 25 suprathreshold samples, shaded orange, is the walking bout. **B.** The raw head yaw angle over the pre-initiation epoch; shading indicates direction relative to straight ahead. **C.** The frame-wise absolute change in yaw. **D.** The running cumulative sum of that change; its value at step-off is that trial’s pre-initiation cumulative absolute head yaw. Abbreviations: Ψscan, cumulative absolute head yaw.

#### Pre–post scanning index

The division of head scanning between the pre-initiation epoch and the walking bout was quantified for each trial as (Ψ_post_ − Ψ_pre_)/(Ψ_post_ + Ψ_pre_), which ranges from −1 (head scanning only in the pre-initiation epoch) to +1 (head scanning only in the walking bout). Because both terms depend on the durations of the pre-initiation epoch and the walking bout, we computed a rate-normalized pre–post scanning index in the same way from the two scanning rates as a sensitivity analysis.

### Statistical analysis

Analyses were performed in MATLAB R2025b (MathWorks, Natick, MA, USA). Each log-transformed outcome was analyzed with a linear mixed-effects model ^21^ (fitlme), log(outcome) ∼ viewing condition × complexity level + (1 | participant). Fixed effects were tested with Type III *F* tests using Satterthwaite degrees of freedom (ANOVA; **Table 2**). The random intercept was grouped by participant, so each participant entered the model once. Pairwise comparisons between viewing conditions were estimated by refitting the model to each pair of conditions with additive fixed effects. Results are reported as ratios of geometric means, and a Bonferroni correction was applied across the three comparisons per outcome (**Table 3**). Pearson correlations across trials related cumulative absolute head yaw to the duration of the corresponding epoch. Both pre–post scanning indices were averaged within participants at low and at high demand, and the two averages were compared within each viewing condition with paired *t*-tests, Bonferroni-corrected across the three viewing conditions, using Cohen’s *d* (mean difference divided by the standard deviation of the differences) as the effect size. Other tests used α = 0.05. Descriptive statistics are medians with median absolute deviations (*MAD*), except for the pre–post scanning index (mean, *SD*).

**Table 2.** Omnibus mixed-effects model terms for every outcome, over the whole trial, in the pre-initiation epoch and in the walking bout.

| **Row** | **Outcome** | **Effect** | ***F*** | ***p*** | **η<sup>2</sup>p** |
|---|---|---|---|---|---|
|  | **Whole trial** |  |  |  |  |
| 1 | Cumulative yaw | Viewing condition | *F*<sub>(2, 37.9)</sub> = 3.79 | 0.032 | 0.167 |
| 2 | Cumulative yaw | Complexity level | *F*<sub>(3, 115.4)</sub> = 58.96 | < 0.001 | 0.605 |
| 3 | Cumulative yaw | Condition × complexity | *F*<sub>(6, 115.4)</sub> = 0.52 | 0.790 | 0.026 |
|  | **Pre-initiation** |  |  |  |  |
| 4 | Cumulative yaw | Viewing condition | *F*<sub>(2, 36.9)</sub> = 4.37 | 0.020 | 0.192 |
| 5 | Cumulative yaw | Complexity level | *F*<sub>(3, 115.8)</sub> = 37.16 | < 0.001 | 0.491 |
| 6 | Cumulative yaw | Condition × complexity | *F*<sub>(6, 115.8)</sub> = 0.88 | 0.514 | 0.044 |
| 7 | Latency to step-off | Viewing condition | *F*<sub>(2, 36.2)</sub> = 11.08 | < 0.001 | 0.380 |
| 8 | Latency to step-off | Complexity level | *F*<sub>(3, 113.9)</sub> = 30.23 | < 0.001 | 0.443 |
| 9 | Latency to step-off | Condition × complexity | *F*<sub>(6, 113.9)</sub> = 0.47 | 0.827 | 0.024 |
| 10 | Scanning rate | Viewing condition | *F*<sub>(2, 37.4)</sub> = 2.24 | 0.120 | 0.107 |
| 11 | Scanning rate | Complexity level | *F*<sub>(3, 115.7)</sub> = 14.17 | < 0.001 | 0.269 |
| 12 | Scanning rate | Condition × complexity | *F*<sub>(6, 115.7)</sub> = 0.66 | 0.684 | 0.033 |
|  | **Walking bout** |  |  |  |  |
| 13 | Cumulative yaw | Viewing condition | *F*<sub>(2, 42.8)</sub> = 0.11 | 0.897 | 0.005 |
| 14 | Cumulative yaw | Complexity level | *F*<sub>(3, 109.2)</sub> = 31.11 | < 0.001 | 0.461 |
| 15 | Cumulative yaw | Condition × complexity | *F*<sub>(6, 109.3)</sub> = 5.23 | < 0.001 | 0.223 |
| 16 | Walking-bout duration | Viewing condition | *F*<sub>(2, 127)</sub> = 3.85 | 0.024 | 0.057 |
| 17 | Walking-bout duration | Complexity level | *F*<sub>(3, 127)</sub> = 8.86 | < 0.001 | 0.173 |
| 18 | Walking-bout duration | Condition × complexity | *F*<sub>(6, 127)</sub> = 0.69 | 0.659 | 0.032 |
| 19 | Scanning rate | Viewing condition | *F*<sub>(2, 40.1)</sub> = 0.35 | 0.707 | 0.017 |
| 20 | Scanning rate | Complexity level | *F*<sub>(3, 111.8)</sub> = 88.56 | < 0.001 | 0.704 |
| 21 | Scanning rate | Condition × complexity | *F*<sub>(6, 111.8)</sub> = 6.68 | < 0.001 | 0.264 |

**Table 3.**
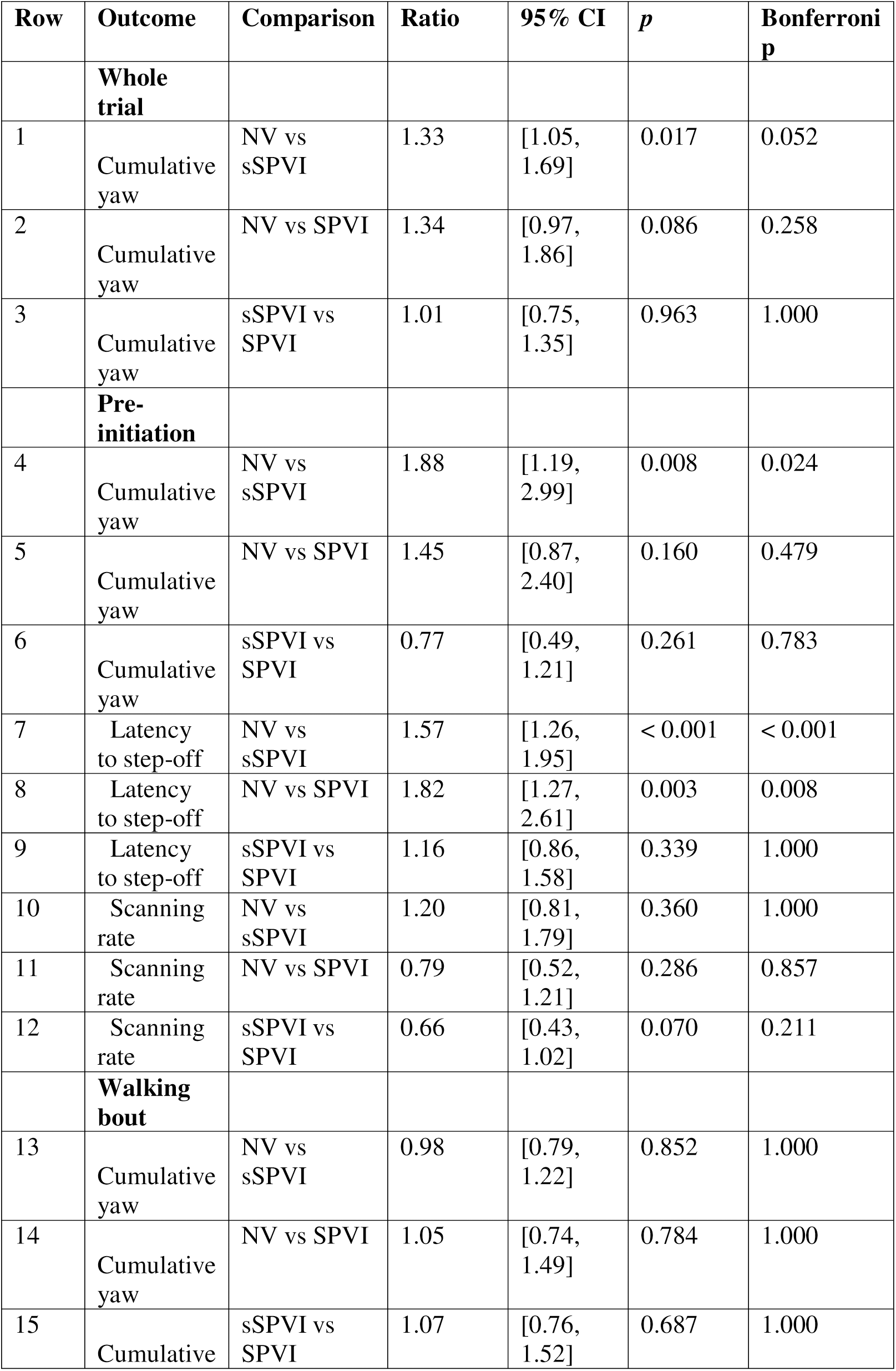
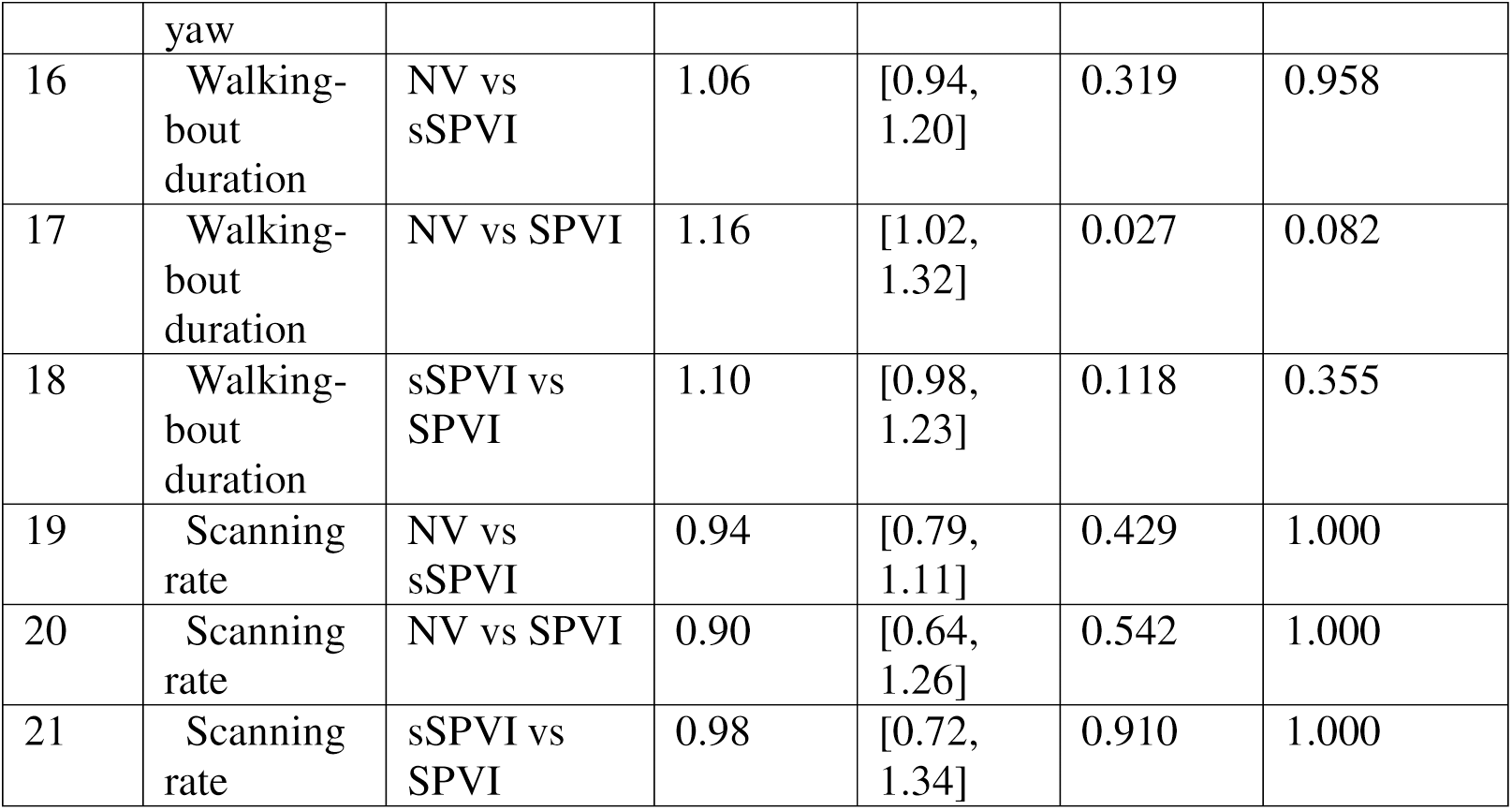
Pairwise comparisons between viewing conditions, from mixed-effects models refitted to each pair of conditions with additive fixed effects; each ratio is the second-named condition relative to the first.

| Row | Outcome | Comparison | Ratio | 95% CI | *p* | Bonferroni p |
|---|---|---|---|---|---|---|
|  | **Whole trial** |  |  |  |  |  |
| 1 | Cumulative yaw | NV vs sSPVI | 1.33 | [1.05, 1.69] | 0.017 | 0.052 |
| 2 | Cumulative yaw | NV vs SPVI | 1.34 | [0.97, 1.86] | 0.086 | 0.258 |
| 3 | Cumulative yaw | sSPVI vs SPVI | 1.01 | [0.75, 1.35] | 0.963 | 1.000 |
|  | **Pre-initiation** |  |  |  |  |  |
| 4 | Cumulative yaw | NV vs sSPVI | 1.88 | [1.19, 2.99] | 0.008 | 0.024 |
| 5 | Cumulative yaw | NV vs SPVI | 1.45 | [0.87, 2.40] | 0.160 | 0.479 |
| 6 | Cumulative yaw | sSPVI vs SPVI | 0.77 | [0.49, 1.21] | 0.261 | 0.783 |
| 7 | Latency to step-off | NV vs sSPVI | 1.57 | [1.26, 1.95] | < 0.001 | < 0.001 |
| 8 | Latency to step-off | NV vs SPVI | 1.82 | [1.27, 2.61] | 0.003 | 0.008 |
| 9 | Latency to step-off | sSPVI vs SPVI | 1.16 | [0.86, 1.58] | 0.339 | 1.000 |
| 10 | Scanning rate | NV vs sSPVI | 1.20 | [0.81, 1.79] | 0.360 | 1.000 |
| 11 | Scanning rate | NV vs SPVI | 0.79 | [0.52, 1.21] | 0.286 | 0.857 |
| 12 | Scanning rate | sSPVI vs SPVI | 0.66 | [0.43, 1.02] | 0.070 | 0.211 |
|  | **Walking bout** |  |  |  |  |  |
| 13 | Cumulative yaw | NV vs sSPVI | 0.98 | [0.79, 1.22] | 0.852 | 1.000 |
| 14 | Cumulative yaw | NV vs SPVI | 1.05 | [0.74, 1.49] | 0.784 | 1.000 |
| 15 | Cumulative yaw | sSPVI vs SPVI | 1.07 | [0.76, 1.52] | 0.687 | 1.000 |
| 16 | Walking-bout duration | NV vs sSPVI | 1.06 | [0.94, 1.20] | 0.319 | 0.958 |
| 17 | Walking-bout duration | NV vs SPVI | 1.16 | [1.02, 1.32] | 0.027 | 0.082 |
| 18 | Walking-bout duration | sSPVI vs SPVI | 1.10 | [0.98, 1.23] | 0.118 | 0.355 |
| 19 | Scanning rate | NV vs sSPVI | 0.94 | [0.79, 1.11] | 0.429 | 1.000 |
| 20 | Scanning rate | NV vs SPVI | 0.90 | [0.64, 1.26] | 0.542 | 1.000 |
| 21 | Scanning rate | sSPVI vs SPVI | 0.98 | [0.72, 1.34] | 0.910 | 1.000 |

#### Ex-Gaussian model of latency to step-off

Because a mean response time does not distinguish a shift of the distribution from a longer slow tail ^14^, latencies to step-off were fitted by maximum likelihood (fminsearch, best of seven starting values) using an ex-Gaussian distribution, the sum of a Gaussian and an exponential component. The Gaussian mean μ and standard deviation σ describe the position and spread of the peak of the latency distribution, and the exponential mean τ describes its slow tail: a larger μ means that every crossing is delayed by a similar amount, whereas a larger τ means that some crossings are delayed much more than others; the mean of the distribution is μ + τ. In attention-deficit/hyperactivity disorder, an elevated τ has been attributed to problems in arousal and state regulation and to difficulty detecting signal from noise ^15^, and in healthy adults, τ is inversely associated with resting heart rate variability ^22^, although no physiological measures were recorded here. Because latencies to step-off were approximately twice as long at high as at low demand (**Figure S2**), pooling the four levels would combine two distributions and increase the estimate of τ; the model was therefore fitted separately at low and at high demand in each viewing condition. Confidence intervals and two-sided *p*-values were obtained from 10,000 refits that resampled participants rather than trials, so that trials of the same participant were not treated as independent ^23^, with the 2.5th and 97.5th percentiles of the resampled distribution as bounds ^24,25^. Comparisons between low and high demand were paired within participants, and comparisons between viewing conditions were unpaired, with Bonferroni correction within each family of three comparisons. σ reached its lower bound of one sampling interval (0.11 s) in four of the six fits and was not compared. Goodness of fit was assessed with Kolmogorov–Smirnov tests.

## RESULTS

### Effects of environmental complexity on outcome measures

The mixed-effects model terms for every outcome are listed in **Table 2**, and the pairwise comparisons between viewing conditions in **Table 3**. Cumulative absolute head yaw over the entire trial increased with complexity, *F*_(3, 115.4)_ = 58.96, *p* < 0.001, from medians of 139° (*MAD* = 75°) and 136° (*MAD* = 68°) at levels A and B to 424° (*MAD* = 120°) and 387° (*MAD* = 127°) at levels C and D. Viewing condition also had a significant effect, *F*_(2, 37.9)_ = 3.79, *p* = 0.032 (NV: *Mdn* = 255°, *MAD* = 148°; sSPVI: *Mdn* = 301°, *MAD* = 162°; SPVI: *Mdn* = 256°, *MAD* = 118°), although no pairwise comparison remained significant after Bonferroni correction (**Table 3, rows 1–3**), and the interaction between viewing condition and complexity was not significant (**Table 2, row 3**).

Cumulative absolute head yaw over time is shown in **Figure 4**, and the means of the three pre-initiation measures at each complexity level in **Figure 5A–C**. In the pre-initiation epoch, cumulative absolute head yaw also increased with complexity. Pooled across viewing conditions, its median was 28° (*MAD* = 18°) at level A, 22° (*MAD* = 15°) at level B, 182° (*MAD* = 66°) at level C and 133° (*MAD* = 64°) at level D. Latency to step-off increased similarly, with medians of 2.71 s (*MAD* = 0.82 s) at level A, 2.95 s (*MAD* = 1.19 s) at level B, 5.53 s (*MAD* = 1.11 s) at level C and 5.71 s (*MAD* = 1.42 s) at level D. Scanning rate also increased, from 8.1 °/s (*MAD* = 4.3 °/s) and 7.4 °/s (*MAD* = 3.8 °/s) at levels A and B to 26.2 °/s (*MAD* = 9.5 °/s) and 22.9 °/s (*MAD* = 10.4 °/s) at levels C and D. From the simplest to the most complex level, cumulative absolute head yaw in the pre-initiation epoch increased 4.70-fold, latency to step-off 2.11-fold, and scanning rate 2.83-fold.

**Figure 4.**
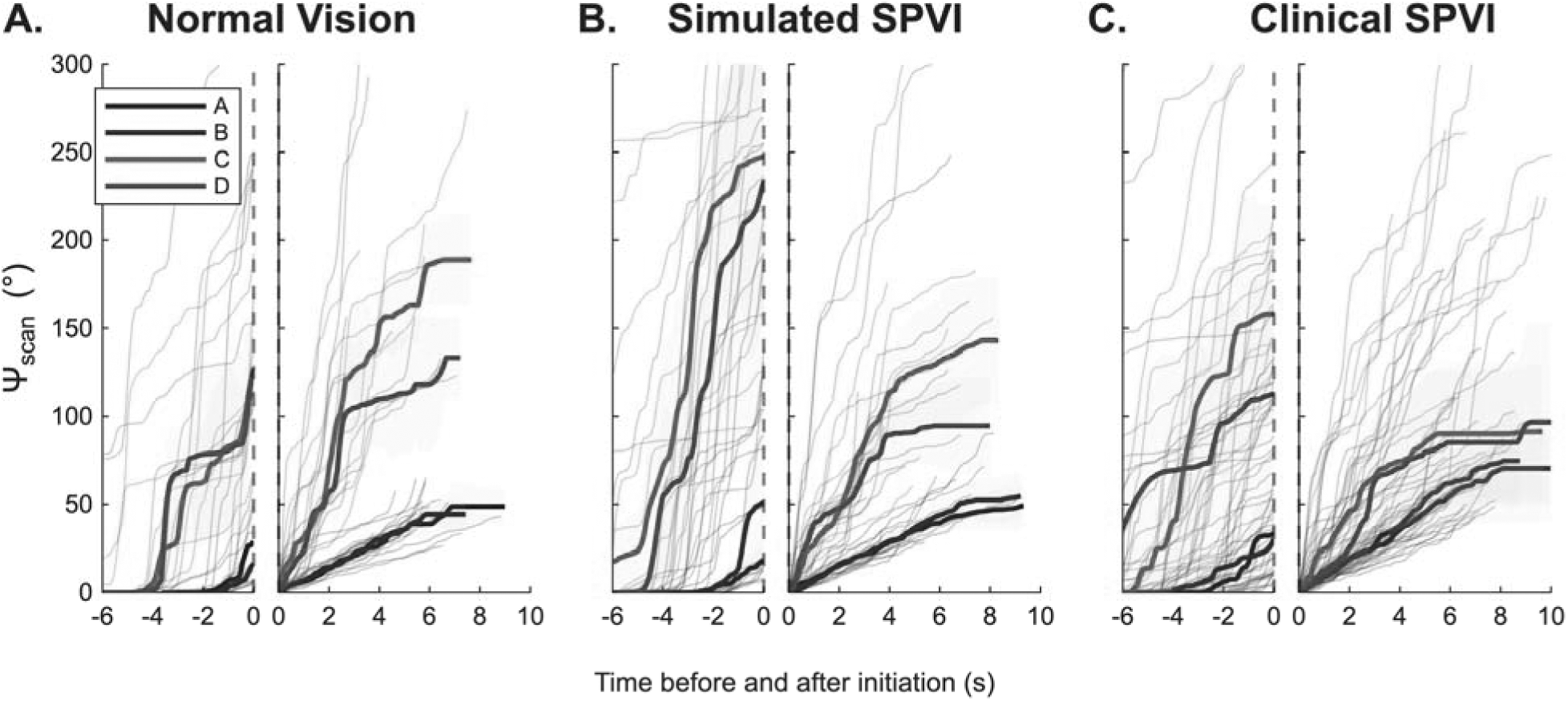
Cumulative absolute head yaw over time, before and during the crossing, by viewing condition and complexity level. Each of the three panels shows one viewing condition, **A.** normal vision, **B.** simulated SPVI, **C.** clinical SPVI, as a pair of plots. The left plot of each pair shows the pre-initiation epoch, referenced to step-off at 0 s; the right plot the first sustained walking bout, referenced to bout onset at 0 s. Thin lines are individual trials. Bold lines are the across-trial median at each time point, colored by complexity level as shown in the legend, and the shaded band spans the median plus and minus one median absolute deviation. The y-axis is capped at 300°; traces exceeding this limit are clipped. The pre-initiation plots include all 144 trials and the walking plots the 139 trials with a qualifying bout. Abbreviations: NV, normal vision; sSPVI, simulated SPVI; SPVI, severe-to-profound visual impairment; Ψscan, cumulative absolute head yaw.

**Figure 5.**
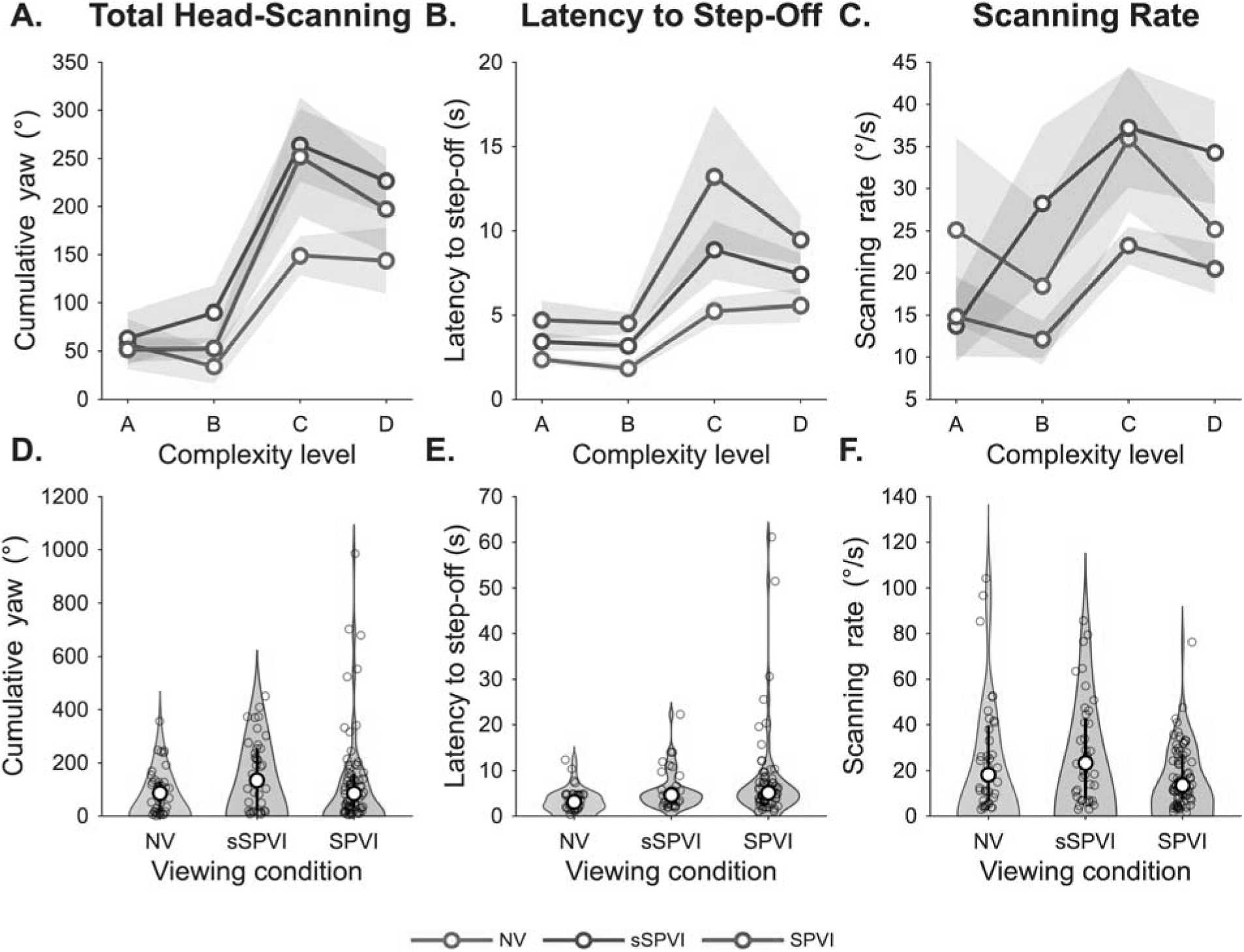
Pre-initiation head scanning by viewing condition and complexity level. The three measures computed over the interval spent standing at the curb: **A, D.** cumulative absolute head yaw; **B, E.** latency to step-off; **C, F.** scanning rate, cumulative yaw divided by latency. In the upper row each symbol is the mean of the contributing trials at one complexity level within one viewing condition and the shaded band spans ± 1 SEM. In the lower row the same measure is pooled across the four complexity levels and shown as a kernel-density violin of the trial distribution, with every trial as a hollow black circle, the interquartile range as a vertical black line, and the median as the white marker. All values are plotted as measured, on linear axes. All 144 trials are included (NV 36, sSPVI 40, SPVI 68).

The mixed-effects models of the pre-initiation epoch are summarized in **Table 2, rows 4–12**. Complexity had a significant main effect on all three measures: cumulative absolute head yaw, *F*_(3, 115.8)_ = 37.16, *p* < 0.001; latency to step-off, *F*_(3, 113.9)_ = 30.23, *p* < 0.001; and scanning rate, *F*_(3, 115.7)_ = 14.17, *p* < 0.001. Viewing condition had a significant main effect on cumulative absolute head yaw, *F*_(2, 36.9)_ = 4.37, *p* = 0.020, and latency to step-off, *F*_(2, 36.2)_ = 11.08, *p* < 0.001, but not on scanning rate (**Table 2, row 10**). The interaction of viewing condition and complexity was not significant for cumulative absolute head yaw, latency to step-off or scanning rate (**Table 2, rows 6, 9 and 12**).

### Differences between viewing conditions before initiation

The comparisons of latency to step-off between viewing conditions are listed in **Table 3, rows 7–9**, and the trial distributions are shown in **Figure 5E**. Compared with normal vision, latency to step-off was 1.57 times longer under simulated SPVI (95% CI [1.26, 1.95], Bonferroni *p* < 0.001) and 1.82 times longer in clinical SPVI (95% CI [1.27, 2.61], Bonferroni *p* = 0.008). Simulated and clinical SPVI did not differ significantly (**Table 3, row 9**).

The corresponding comparisons for cumulative absolute head yaw and scanning rate are listed in **Table 3, rows 4–6 and 10–12**, and the trial distributions are shown in **Figure 5D and F**. In the pre-initiation epoch, cumulative absolute head yaw was 1.88 times higher under simulated SPVI than under the participants’ own normal vision (95% CI [1.19, 2.99], Bonferroni *p* = 0.024). Clinical SPVI did not differ significantly from normal vision in cumulative absolute head yaw (**Table 3, row 5**) or in scanning rate (**Table 3, row 11**).

### During the walking bout

Head scanning in the walking bout is shown in **Figures 4 and 6**, and the corresponding model terms are listed in **Table 2, rows 13–21**. In the walking bout, complexity again had a significant effect on cumulative absolute head yaw, *F*_(3, 109.2)_ = 31.11, *p* < 0.001, and scanning rate, *F*_(3, 111.8)_ = 88.56, *p* < 0.001, but this effect depended on viewing condition, with significant interactions for cumulative absolute head yaw, *F*_(6, 109.3)_ = 5.23, *p* < 0.001, and scanning rate, *F*_(6, 111.8)_ = 6.68, *p* < 0.001. The interaction between viewing condition and complexity was attributable to a steep increase in cumulative absolute head yaw with complexity under NV and sSPVI. In NV, the median cumulative absolute head yaw in the walking bout was 49° (*MAD* = 10°) at level A and 44° (*MAD* = 8°) at level B, and 189° (*MAD* = 26°) at level C and 133° (*MAD* = 23°) at level D. In sSPVI, cumulative absolute head yaw increased similarly, from 49° (*MAD* = 13°) and 55° (*MAD* = 13°) to 143° (*MAD* = 36°) and 95° (*MAD* = 28°). In SPVI, cumulative absolute head yaw changed comparatively little: 75° (*MAD* = 26°), 71° (*MAD* = 21°), 91° (*MAD* = 38°), and 97° (*MAD* = 57°). Scanning rate in the walking bout also increased with complexity in every viewing condition, but to different extents.

**Figure 6.**
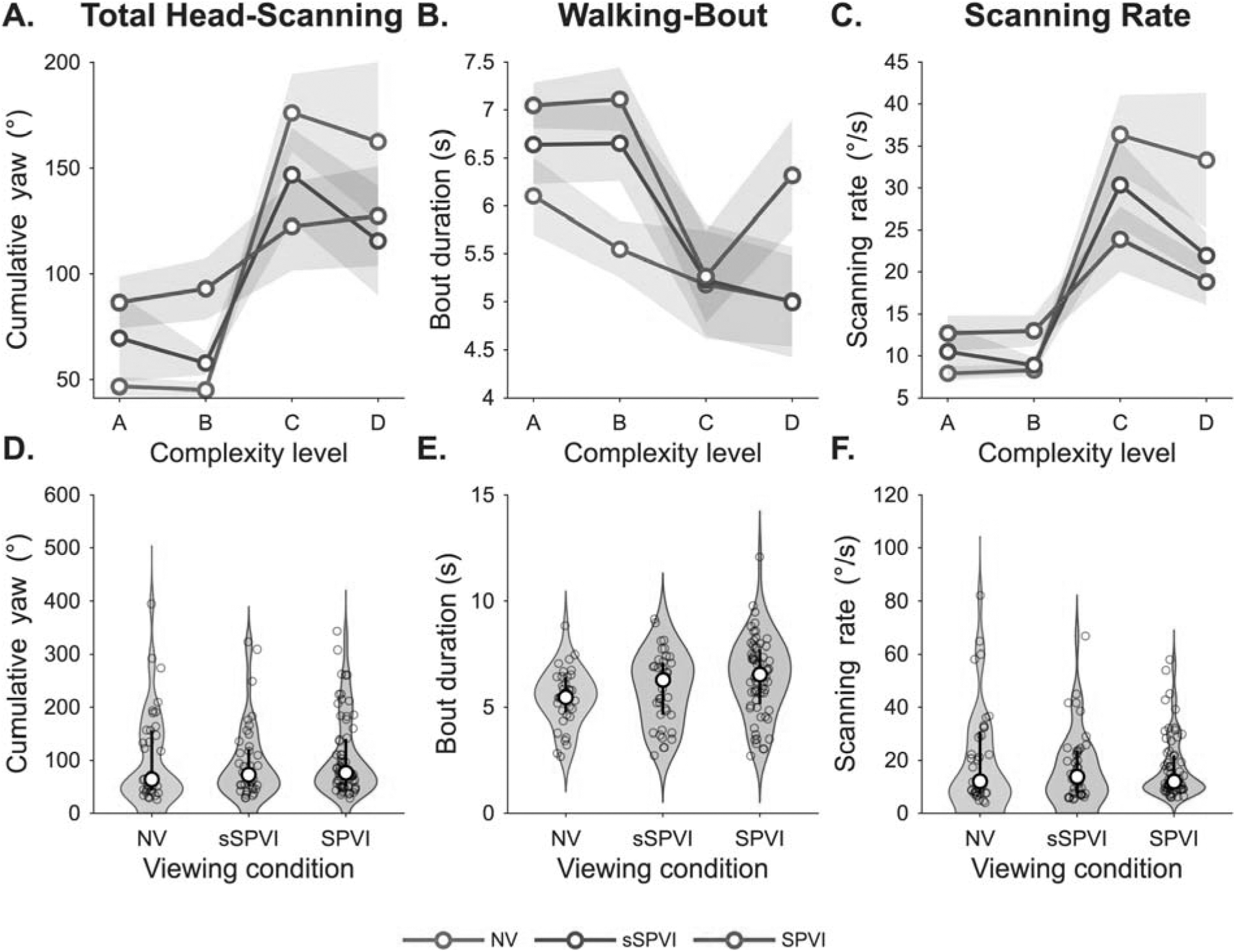
Head scanning during the first sustained walking bout, by viewing condition and complexity level. Same layout as Figure 5, for the walking bout: **A, D.** cumulative absolute head yaw; **B, E.** walking-bout duration; **C, F.** scanning rate, cumulative yaw divided by bout duration. Upper row, mean with a ± 1 SEM band; lower row, kernel-density violin of the pooled trial distribution, with every trial as a hollow black circle, the interquartile range as a vertical black line, and the median as the white marker. All values are plotted as measured, on linear axes. Only the 139 trials containing a qualifying walking bout are included (NV 36, sSPVI 40, SPVI 63).

The effects of viewing condition in the walking bout are listed in **Tables 2 and3, rows 13–21**. Viewing condition had no significant main effect on cumulative absolute head yaw in the walking bout (**Table 2, row 13**) or scanning rate (**Table 2, row 19**). No pairwise comparison of either cumulative absolute head yaw or scanning rate approached significance in the walking bout (**Table 3, rows 13–15 and 19–21**).

Walking-bout duration is shown in **Figure 6B and E**, and its model terms and comparisons are listed in **Tables 2 and3, rows 16–18**. Walking-bout duration was also affected by complexity, *F*_(3, 127)_ = 8.86, *p* < 0.001, with shorter bouts at levels C and D (*Mdn* = 5.04 s, *MAD* = 1.40 s, and *Mdn* = 5.59 s, *MAD* = 1.73 s) than at levels A and B (*Mdn* = 6.60 s, *MAD* = 0.79 s, and *Mdn* = 6.58 s, *MAD* = 1.11 s), and by viewing condition, *F*_(2, 127)_ = 3.85, *p* = 0.024, but not by their interaction (**Table 2, row 18**). For this outcome, the participant variance was estimated at the boundary of zero, so the random intercept was not identifiable and residual denominator degrees of freedom are reported. Walking bouts were longer in SPVI than in NV, but this comparison of walking-bout duration was not significant after Bonferroni correction (**Table 3, row 17**). Simulated SPVI did not differ significantly from either normal vision (**Table 3, row 16**) or clinical SPVI (**Table 3, row 18**).

Cumulative absolute head yaw is plotted against epoch duration in **Figure 7**, for the pre-initiation epoch (A) and the walking bout (B). Across all 144 trials, cumulative absolute head yaw in the pre-initiation epoch and latency to step-off were strongly correlated, *r*_(142)_ = 0.70, *p* < 0.001, whereas cumulative absolute head yaw in the walking bout and walking-bout duration were only weakly correlated, *r*_(137)_ = 0.19, *p* = 0.027. Walking-bout duration spanned a narrower range than latency to step-off (2.69–12.07 s vs. 0.32–61.10 s).

**Figure 7.**
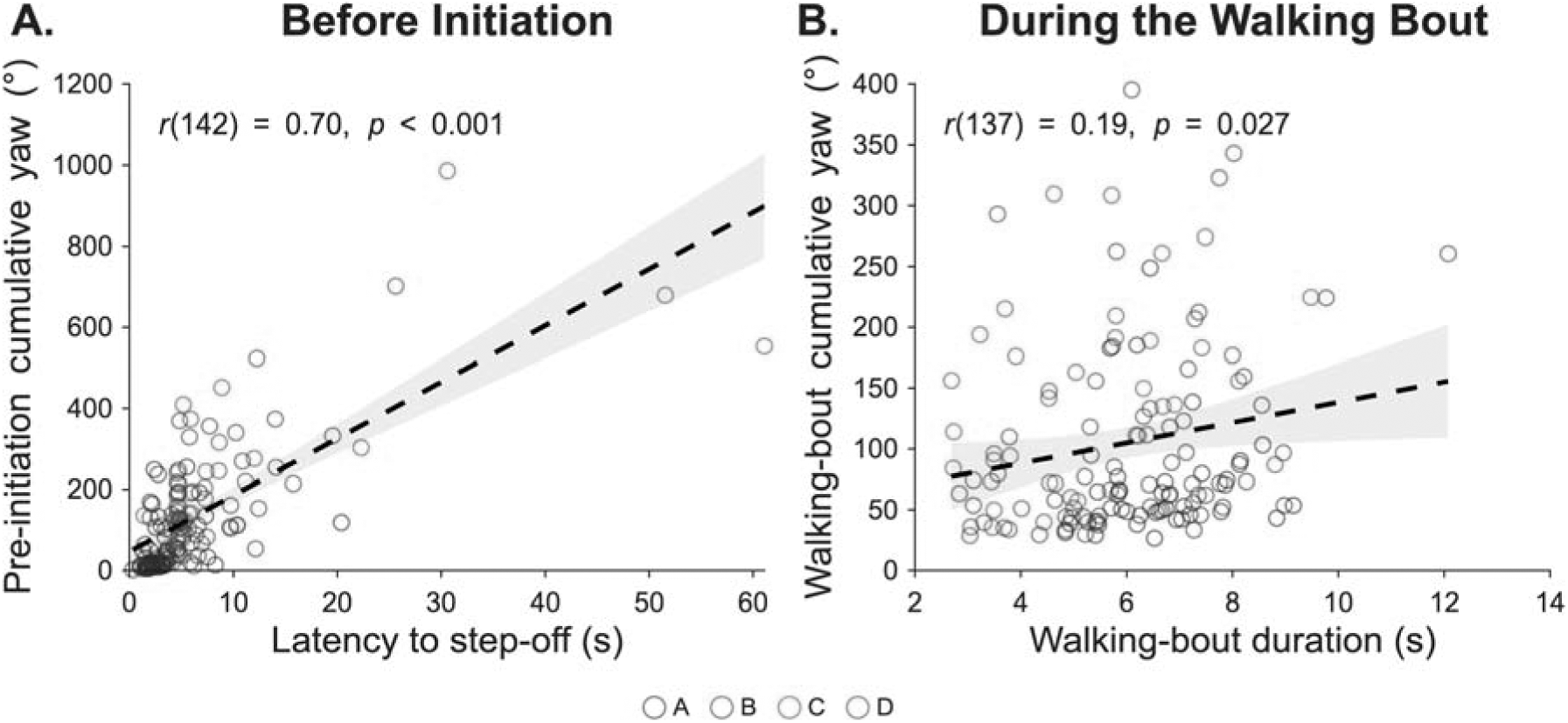
Relationship between scanning and epoch duration, before and during the crossing. **A.** Pre-initiation cumulative absolute head yaw against latency to step-off, one point per trial, all 144 trials. **B.** Walking-bout cumulative absolute head yaw against walking-bout duration, one point per trial, for the 139 trials with a qualifying bout. Points are colored by complexity level (A to D) as shown in the legend. The dashed line is the ordinary least-squares fit and the gray band its 95% confidence interval. Each panel reports the Pearson correlation computed on the trials shown.

### Pre–post scanning index

The pre–post scanning index is shown for each complexity level in **Figure 8A** and averaged at low and at high demand in **Figure 8B**. It was positive in the two simplest environments and near zero or negative in the two most complex ones. Pooled over complexity, its mean was 0.17 (*SD* = 0.42) for NV, −0.06 (*SD* = 0.47) for sSPVI and 0.09 (*SD* = 0.51) for SPVI.

**Figure 8.**
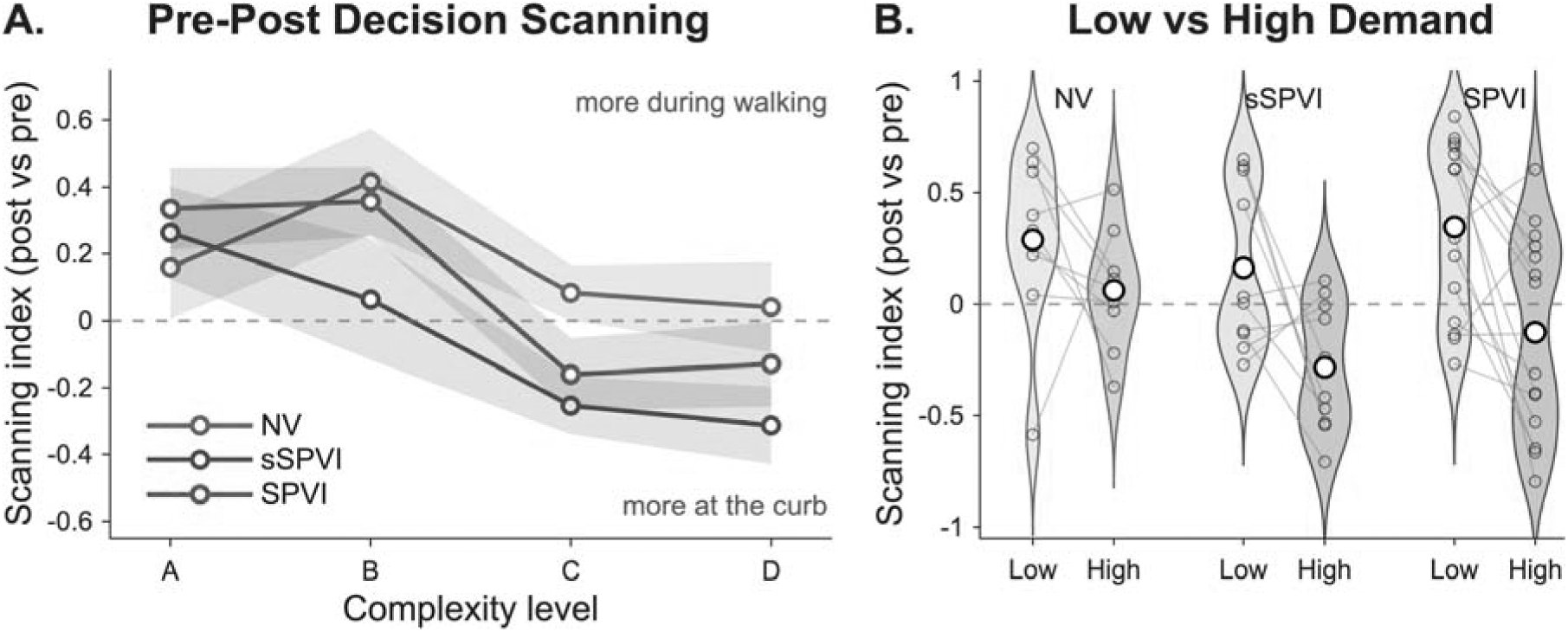
Pre–post scanning index by complexity level and viewing condition. The pre–post scanning index is (post − pre) / (post + pre), where post and pre are cumulative absolute head yaw during the first walking bout and before step-off. Positive values indicate proportionally more scanning during the crossing, negative values proportionally more scanning before it, and the dashed horizontal line at 0 an equal division between the pre-initiation epoch and the walking bout. **A.** The pre–post scanning index across the four complexity levels, one line per viewing condition; symbols are the trial-level mean and the shaded band spans ± 1 SEM. **B.** Each participant’s mean pre–post scanning index over levels A and B (low demand) and over levels C and D (high demand), joined by a gray line and shown as hollow black circles over a kernel-density violin of the participant means; the white marker is the group mean for each pair of levels. Panel A is based on 36 NV, 40 sSPVI and 63 SPVI trials; panel B on 9, 10 and 15 participants.

From low to high demand (**Figure 8B**), the pre–post scanning index decreased significantly in SPVI, from *M* = 0.35 (*SD* = 0.38) to *M* = −0.13 (*SD* = 0.44), *t*_(14)_ = 3.59, Bonferroni *p* = 0.009, *d* = 0.93 (*N* = 15 of 17; two participants had no usable index in one of the two demand conditions). In sSPVI, the pre–post scanning index decreased by a similar amount, from *M* = 0.16 (*SD* = 0.37) to *M* = −0.28 (*SD* = 0.29), but not significantly after Bonferroni correction, *t*_(9)_ = 2.48, Bonferroni *p* = 0.105, *d* = 0.78, *N* = 10. The pre–post scanning index did not change significantly in NV, from *M* = 0.29 (*SD* = 0.39) to *M* = 0.06 (*SD* = 0.27), *t*_(8)_ = 1.25, Bonferroni *p* = 0.743, *d* = 0.42, *N* = 9. For the rate-normalized pre–post scanning index, the change from low to high demand was not significant in any viewing condition (all Bonferroni *p* ≥ 0.603) and reversed sign under normal vision.

### Modeling latency to step-off

The ex-Gaussian fits are shown in **Figure 9**, with the fitted densities in panels A and B and the parameters in panels F and G. At low demand, τ increased with vision loss: 0.18 s in NV, 1.48 s in sSPVI and 3.58 s in SPVI. Only the comparison between NV and SPVI was significant, with τ 3.40 s longer in SPVI (95% CI [0.89, 4.75], Bonferroni *p* = 0.037). SPVI did not differ significantly from sSPVI (2.10 s, 95% CI [−0.01, 3.97], Bonferroni *p* = 0.153), nor sSPVI from NV (1.30 s, 95% CI [−0.31, 1.86], Bonferroni *p* = 0.325). At high demand, τ was longer in every viewing condition: 2.12 s in NV, 4.10 s in sSPVI and 8.32 s in SPVI. Again, only the comparison between NV and SPVI was significant (6.20 s, 95% CI [1.42, 11.35], Bonferroni *p* = 0.022), whereas SPVI did not differ significantly from sSPVI (4.22 s, 95% CI [−0.76, 9.12], Bonferroni *p* = 0.315). At both low and high demand, τ in sSPVI was intermediate between NV and SPVI and did not differ significantly from either.

**Figure 9.**
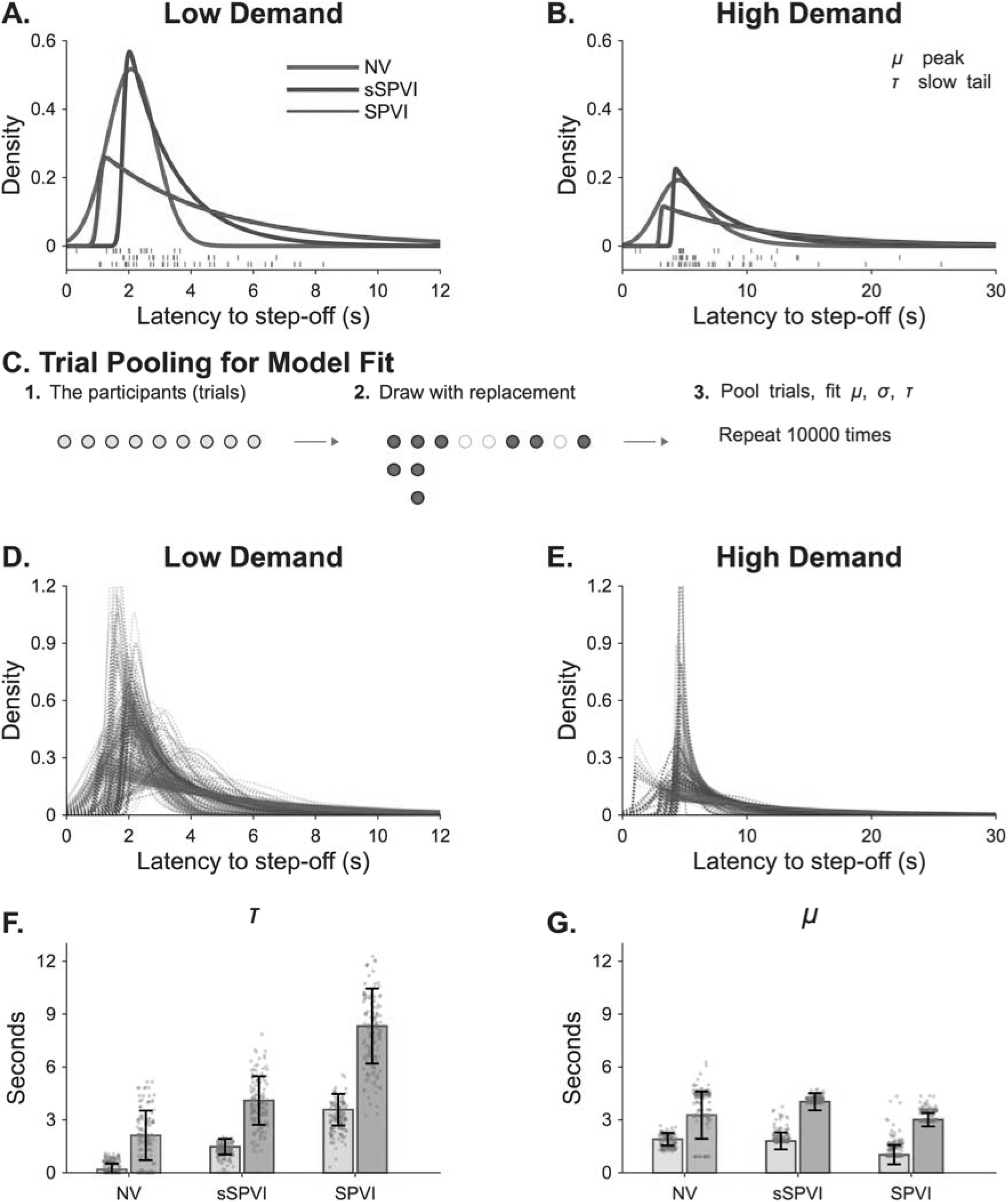
Ex-Gaussian decomposition of latency to step-off, fitted separately to levels A and B (low demand), which contained no moving vehicles, and to levels C and D (high demand), which contained moving vehicles. **A, B.** The fitted densities for levels A and B and for levels C and D, with individual trials as ticks below the axis, one row per viewing condition. The key in B defines μ, the position of the peak of the latency distribution, and τ, the slow tail of the latency distribution. C. Participant-level bootstrap: participants are resampled with replacement (stacked filled circles, participants drawn more than once; empty rings, participants not drawn), all trials of a resampled participant are retained together, and the model is refitted. D, E. Densities of 100 of the 10,000 refits for each viewing condition, on a taller vertical scale than in A and B. F. The exponential component τ and **G.** the Gaussian component μ, in each viewing condition, with levels A and B in the lighter bar and levels C and D in the darker bar. Bar height is the fit to the observed data; error bars are ±1 standard error, computed as the standard deviation of the refits and truncated at zero because neither parameter can be negative. Both panels share a common vertical scale in seconds. One hundred and fifty refits are overlaid as small gray dots. σ is not plotted. Each fit uses 18 (NV), 20 (sSPVI) or 34 (SPVI) trials per pair of levels. Abbreviations: μ, mean of the Gaussian component; σ, standard deviation of the Gaussian component; τ, mean of the exponential component.

The position of the peak of the latency distribution, μ, is shown in **Figure 9G**, and the cumulative distributions of latency to step-off in **Figure S2**. Unlike τ, μ did not differ significantly between any two viewing conditions at low or at high demand (all Bonferroni *p* ≥ 0.256). At low demand, μ was 1.90 s in NV, 1.81 s in sSPVI and 1.02 s in SPVI, a decrease with vision loss, opposite in direction to the difference in latency to step-off; at high demand, μ was 3.27, 4.03 and 3.01 s, with no consistent order with vision loss. Accordingly, the fastest SPVI crossings were close to those under normal vision (10th percentile 1.59 vs 1.35 s at low demand), whereas 76% of SPVI crossings (sSPVI, 90%) exceeded the median NV latency to step-off and 47% (sSPVI, 25%) exceeded the latency to step-off of the slowest NV crossing; at high demand, 74% (sSPVI, 75%) exceeded the median NV latency to step-off. All fitted parameters and comparisons are available with the analysis code in the Zenodo deposit.

Both μ and τ were larger at high than at low demand in every viewing condition (**Figure 9F, G**). The increase in μ was of similar magnitude in all three (1.37 s in NV, 2.22 s in sSPVI and 1.98 s in SPVI) and was significant only in sSPVI after Bonferroni correction (Bonferroni *p* = 0.846, 0.025 and 0.107, respectively). The increase in τ was larger with greater vision loss and significant only in SPVI, where τ increased by 4.74 s (95% CI [1.09, 9.19], Bonferroni *p* = 0.034); the increases in NV (1.94 s) and sSPVI (2.62 s) were not significant. The increase in τ from low to high demand did not differ significantly between viewing conditions; the largest difference, between SPVI and NV, was 2.81 s (95% CI [−1.36, 8.48], Bonferroni *p* = 0.498).

## DISCUSSION

### Time rather than scanning

Of the two forms of compensation considered in the Introduction, only additional time at the curb was observed: participants with SPVI took longer to step off than normally sighted participants, whereas cumulative absolute head yaw did not differ significantly between SPVI and normal or simulated vision, either in the pre-initiation epoch or in the walking bout. Because cumulative absolute head yaw accumulates with time, the additional time at the curb would by itself be expected to increase it; its smaller, non-significant increase in SPVI reflects a scanning rate that was, if anything, lower than under normal vision (**Table 3, rows 5 and 11**). These null results for cumulative absolute head yaw have wide confidence intervals and do not demonstrate equivalent head scanning; moreover, they were not consistent across complexity levels: in the walking bout, cumulative absolute head yaw increased steeply with complexity under normal and simulated vision but only slightly in SPVI (**Figure 6A**). Across viewing conditions, head scanning was influenced more by environmental complexity than by vision.

### Compensatory scanning at the severe end of vision loss

The prediction that degraded vision requires more extensive head scanning was not supported at the severe end of vision loss. Compensatory head and eye scanning presupposes sufficient residual form vision for directed head movements to be informative: it has been documented where a defined blind field provides a direction for head and eye scanning, as in hemianopia ^8,9^, and where a useful visual field remains, as in early to moderate retinitis pigmentosa, in which head movement increases at the expense of eye movement, whereas participants with late-stage disease made minimal gaze movements ^11^. With severely degraded vision, the available compensation may instead be additional time. The absence of increased head scanning in SPVI is consistent with reduced or unchanged head turning in macular degeneration ^10^, reduced eye scanning in retinitis pigmentosa with tunnel vision ^26^, and fewer head turns in the combined-loss group closest to SPVI in a study in which patients, judging a recorded scene while stationary, also identified safe gaps later than controls ^13^. Earlier studies measured the accuracy of gap judgments in pedestrians with less severe vision loss or, with hearing alone, in blind pedestrians ^4,5^; we measured latency to step-off on a task that every participant completed. Together, these findings are consistent with the view that vision loss limits the acquisition of information rather than its evaluation, so that degraded input increases the time required; whether accuracy declines when that time is not available was not tested here. The present analysis quantified the extent of head yaw, not its direction.

### Locus of the additional time

In clinical SPVI, the slow tail of the latency distribution was longer than under normal vision at both low and high demand, without a detectable shift of its peak, a dissociation that distributional analyses of response times are designed to detect ^14^. The fastest crossings in SPVI were therefore initiated almost as promptly as under normal vision, whereas most were delayed by a variable rather than a fixed amount. In diffusion-model simulations of speeded two-choice decisions, poorer-quality evidence, expressed as a lower drift rate, results mainly in a larger τ; a longer non-decision time (sensory encoding and motor execution) results only in a larger μ; and greater caution, expressed as a wider decision boundary, results in a larger τ but also in a substantially larger μ ^16,27^. Of the two determinants of the time spent at the curb outlined in the Introduction, a longer slow tail of the latency distribution without a significant shift of its peak is therefore more consistent with slower acquisition of visual information about approaching vehicles than with a larger amount of information required before committing, although μ was imprecisely estimated under the participant-level bootstrap, ex-Gaussian parameters do not map uniquely onto diffusion-model parameters, and the two determinants cannot be fully separated without a measure of accuracy ^16^. If the longer slow tail of the latency distribution reflected attentional lapses, to which an elevated τ has been attributed in attention-deficit/hyperactivity disorder and in healthy adults ^15,22^, only occasional crossings, rather than most, would be expected to be prolonged. The slow tail of the latency distribution was already longer at levels A and B, which contained no moving vehicles, so the effect of vision loss did not depend on traffic; at levels C and D, which contained moving vehicles, the slow tail of the latency distribution was significantly longer than at levels A and B only in SPVI, but this increase did not differ significantly between viewing conditions, so whether traffic adds to the effect of vision loss remains unresolved.

Two alternative explanations are also consistent with the data on latency to step-off. Because the task provided no auditory or tactile traffic information and cane trials were excluded, prolonged latencies to step-off may reflect the absence of visual evidence about approaching vehicles rather than its slow accumulation. In addition, the vehicles at levels C and D yielded, which makes these latencies to step-off incomparable with the gap-acceptance literature; because yielding is likely to be difficult to detect at this level of vision, part of the slow tail of the latency distribution may reflect the time taken to abandon the judgment of whether vehicles had yielded rather than to complete it.

### Acute simulation and long-standing vision loss

Comparing simulated with clinical SPVI separates the immediate effect of acuity loss from adaptation to long-standing vision loss. On mean latency to step-off, the sighted controls with the Bangerter filters showed most of the additional latency to step-off of clinical SPVI, and simulated and clinical SPVI did not differ significantly on any measure (**Table 3, rows 3, 6, 9, 12, 15, 18 and 21**); the additional latency to step-off therefore does not require long-standing vision loss. Cumulative absolute head yaw in the pre-initiation epoch was also higher with the Bangerter filters but not in clinical SPVI; the increase with the filters may reflect a more conservative decision criterion under unfamiliar blur. The smaller increase in cumulative absolute head yaw with complexity in the walking bout in SPVI, which was not observed under simulated SPVI, together with walking bouts that were no shorter than under normal vision, suggests that participants with long-standing vision loss made the decision to step off before initiating crossing and then crossed with little further visual updating of the traffic scene, whereas participants under the acute blur produced by the Bangerter filters continued to scan as sighted pedestrians do; neither difference was tested directly. The slow tail of the latency distribution was longest in SPVI, although acuity with the Bangerter filters was poorer than that of 11 of the 14 participants with SPVI whose acuity was recorded (**Table 1**); any excess over simulation, if confirmed, would therefore reflect aspects of clinical SPVI that the Bangerter filters do not simulate, such as field loss, disease-specific visual characteristics and age, or strategies adapted to non-visual cues. After vision loss, some auditory abilities have been proposed to improve and others to decline ^28^, and pedestrians who are blind rely on hearing to detect approaching vehicles where there is no traffic control ^29^; because no auditory traffic information was presented in the virtual task, participants with long-standing vision loss were tested without the cues on which their adapted strategies depend. The outcome under simulation can therefore match that of clinical vision loss even when the underlying process differs, a caveat for the routine use of acuity simulation, including in our previous work ^19^.

### Distribution of head scanning before and during the crossing

At high complexity, participants with clinical SPVI allocated a larger proportion of their cumulative absolute head yaw to the pre-initiation epoch (**Figure 8**); under simulated SPVI, the shift was of similar size but not significant after correction. Computed from scanning rate, the pre–post scanning index did not change significantly, so the larger proportion reflects a longer pre-initiation epoch rather than a higher scanning rate at the curb relative to that during the walking bout: with moving vehicles, participants with SPVI spent longer at the curb, whereas walking bouts were not longer at higher complexity and their cumulative absolute head yaw in the walking bout increased only slightly. Consistent with this, cumulative absolute head yaw was more strongly correlated with the duration of the pre-initiation epoch than with that of the walking bout (**Figure 7**), although the narrower range of walking-bout durations limits this comparison. The redistribution of head scanning toward the curb is therefore a consequence of the additional time at the curb rather than a separate form of compensation, consistent with a decision to step off that was completed before crossing was initiated.

### Implications for orientation and mobility

The additional time at the curb could reflect slower acquisition of visual information about approaching vehicles or a larger amount of information required before committing, and the two have different implications. If vision loss slows acquisition, as the present results suggest, instructing a patient or client to scan more would not be expected to help, whereas preserving time at the curb would; if it increases the amount of information required, the time spent at the curb reflects a strategy that may be amenable to training. In either case, limited head movement at a curb does not by itself indicate that a patient or client is not surveying the street, and training environments could be graded by environmental complexity rather than by acuity. Both remain hypotheses. To our knowledge, no study has shown that increased head or eye scanning improves crossing safety in low vision; in drivers with hemianopia, auditory reminders increased early head scanning, and early scans were associated with faster responses to hazards in a simulator, but whether this improves safety has not been tested prospectively ^9^. Controlled evidence for orientation and mobility training in low vision is limited ^30,31^, and we are not aware of any trial of street-crossing training in adults with SPVI. Because the additional time at the curb was variable and often long rather than fixed, crossing aids timed to average pedestrian performance could be insufficient for many crossings; whether a vehicle has yielded could instead be conveyed non-visually, for example by a wearable vibrotactile display calibrated to the wearer’s vibrotactile sensitivity ^32^.

### Limitations

Crossing initiation is delayed in virtual environments relative to a real test track, and effects observed in simulators can be absent on the track ^33^, so neither the absolute latencies to step-off nor necessarily their pattern may generalize directly to real streets; differences between age groups, however, were reproduced in both a CAVE simulator and a head-mounted display for most crossing measures ^34^, and conventional orientation and mobility assessment is itself imperfectly standardized ^35^. The ex-Gaussian fits pooled two trials per participant at each pair of levels, far fewer than are typically used for such fits; τ therefore describes the group distribution and cannot distinguish long slow tails of the latency distribution within individuals from consistently slower participants, and the goodness-of-fit test, computed on the same trials, is anti-conservative. With 9, 10 and 17 sessions per condition, the between-condition comparisons had limited statistical power. Among the 14 participants with SPVI whose acuity was recorded, acuity was concentrated between 1.0 and 1.6 logMAR (10 of 14; **Table 1**), too narrow a range to test whether the effects varied with acuity; in an exploratory analysis, acuity was not significantly correlated with latency to step-off (geometric mean per participant), Spearman *r*_(12)_ = 0.27, *p* = 0.347. Head yaw is not equivalent to gaze direction. The virtual environment posed no physical risk; and the complexity levels were presented in a fixed order, so their effects are confounded with practice. Whether head-yaw kinematics predict travel incidents outside the laboratory remains to be determined in prospective studies.

## Conclusions

Pedestrians with SPVI took longer to step off than normally sighted pedestrians, and the additional time was variable and often long rather than fixed; they did not scan at a higher rate, so any increase in their cumulative head scanning resulted from the additional time. When vehicles were moving, a larger share of their head scanning occurred before step-off, because they spent longer at the curb rather than because their scanning rate at the curb rose relative to that during walking. For pedestrians with SPVI, the time spent at the curb, and its variable, right-skewed lengthening, may therefore be a more informative target than the extent of head scanning, both for orientation and mobility assessment and for the design of accessible crossings, particularly uncontrolled crossings at which no non-visual information about yielding vehicles is available.

## Declarations

## Conflict of interest statement

Kartha, Dagnelie and Sadeghi are named inventors on intellectual property managed by Johns Hopkins Technology Ventures; the intellectual property is licensed, and they receive royalties from it. Tran and Gee are affiliated with BaltiVirtual, which developed the virtual environments used in this study. Erkat-Cohen, Venugopal and Livingston declare no competing interests.

## Funding

This work was supported by the National Eye Institute (R00EY033031 to AK; R01EY028452 to GD) and by Research to Prevent Blindness (GD). The funders had no role in study design, data collection and analysis, decision to publish, or preparation of the manuscript.

## Data availability statement

The de-identified trial-level data and the MATLAB code that reproduce every figure, table and statistic are available on Zenodo (https://doi.org/10.5281/zenodo.23955167). The raw head-pose logs are available from the corresponding author on reasonable request.

## Author contributions

Conceptualization: BEC, AK. Methodology: BEC, DV, AK, GD. Software: CT, WG. Formal analysis: BEC, DV. Investigation: BEC, AK. Data curation: AK, RS. Writing, original draft: BEC. Writing, review and editing: all authors. Visualization: BEC. Supervision: AK. Funding acquisition: AK, GD. All authors approved the final version of the manuscript.

## Ethics approval and consent

The study adhered to the tenets of the Declaration of Helsinki. Johns Hopkins Medicine Institutional Review Board approval was obtained. Written informed consent was obtained from the participants after explanation of the study prior to enrollment.

## Acknowledgements

We would like to acknowledge the Multisensory Integration in Visual Impairment Laboratory and SUNY Optometry community for helpful discussions on the topic.

## SUPPLEMENTARY INFORMATION

**Figure S1.**
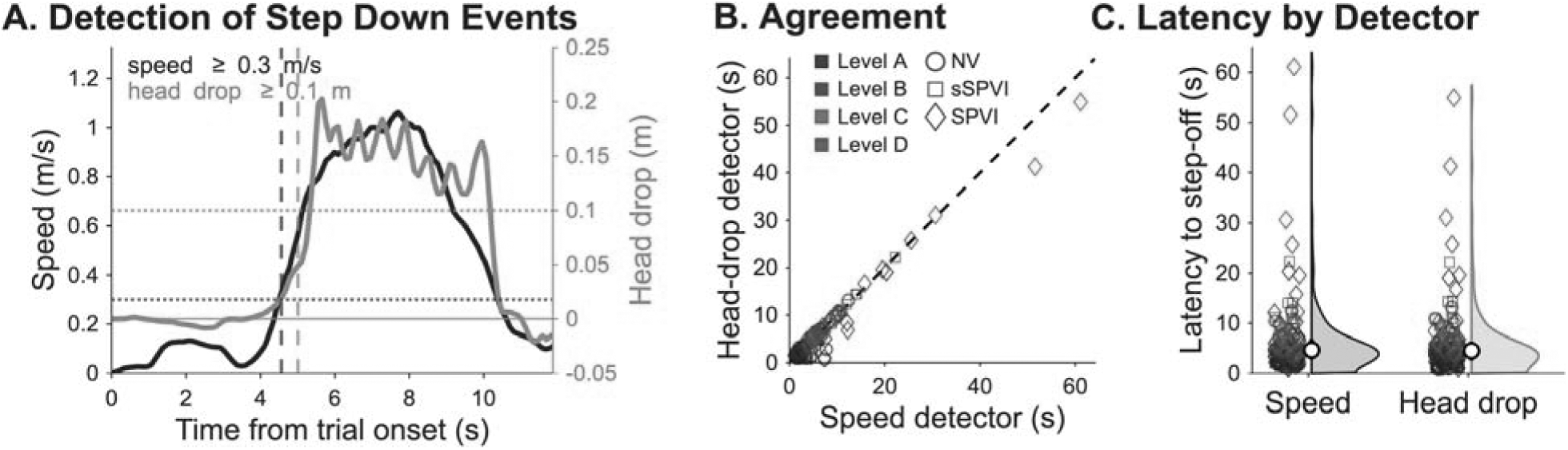
Step-off detection by walking speed and by curb step-down. In this study, step-off is detected when horizontal walking speed reaches 0.3 m/s; our recent work ^20^ detects the same event from a vertical drop of the head of at least 0.1 m within 0.5 m of the starting position. **A.** One trial, showing smoothed walking speed in black against the left axis and the drop in head height in orange against the right axis, with the threshold for each shown as a dotted line and the resulting step-off times as dashed vertical lines. **B.** The two latencies plotted against each other for every trial, with the identity line dashed; they were highly correlated, Spearman *r*_(134)_ = 0.87, *p* < 0.001, and Pearson *r*_(134)_ = 0.98, *p* < 0.001. **C.** Latency to step-off under each detector, pooled over viewing conditions and complexity levels, shown as the individual trials on the left and a half violin of their kernel density on the right, estimated with the same Gaussian kernel for both detectors, with the median as the white marker and the interquartile range as the vertical black line. The two distributions did not differ, two-sample Kolmogorov–Smirnov *D* = 0.10, *p* = 0.446, nor did the paired latencies, Wilcoxon signed-rank *z* = 0.84, *p* = 0.401. Within each viewing condition and complexity level, no combination of viewing condition and complexity level differed significantly (all *D* ≤ 0.50, all *p* ≥ 0.111; all Bonferroni-corrected *p* = 1.000 across the 12 combinations), and the paired signed-rank test gave the same result after Bonferroni correction (smallest corrected *p* = 0.328). In B and C every trial is a hollow marker colored by complexity level and shaped by viewing condition (circle, NV; square, sSPVI; diamond, SPVI). Panels B and C use the 136 trials in which a step-down was detected (NV 33, sSPVI 39, SPVI 64); in the other 8 of the 144 trials no step-down was detected, so a head-drop latency is undefined.

**Figure S2.**
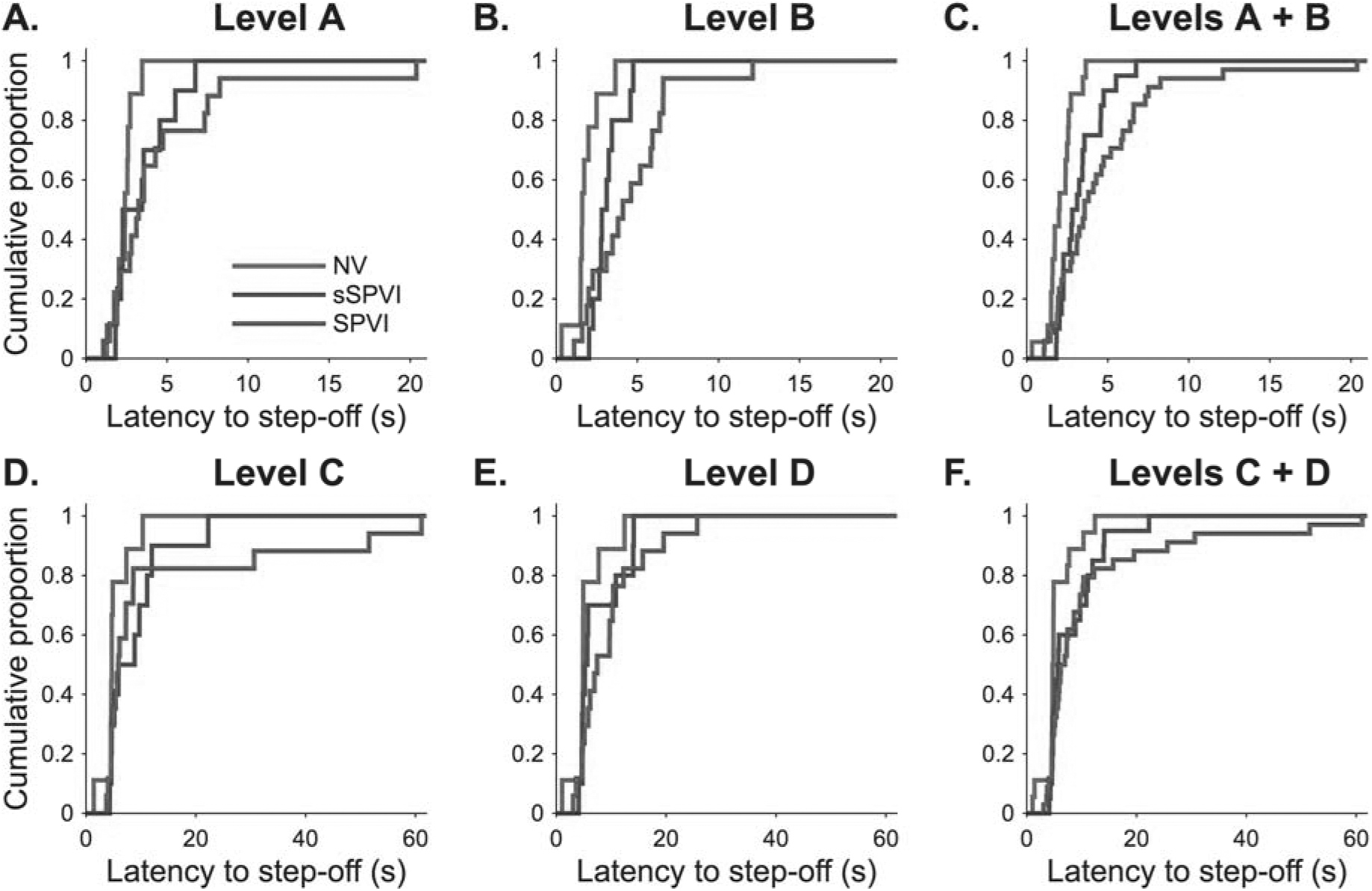
Cumulative distributions of latency to step-off at each complexity level and pooled over the pairs of levels modeled in Figure 9. **A.** Level A, **B.** level B and **C.** levels A and B pooled (low demand); **D.** level C, **E.** level D and **F.** levels C and D pooled (high demand). Each curve is the empirical cumulative distribution of one viewing condition, with no fit overlaid. The panels of each row share a horizontal range spanning the full extent of the data in that row, so every curve reaches 1 and the long slow tail of the latency distribution at levels C and D is visible; the density panels of Figure 9 are plotted over a shorter range containing most of the data, and 2 trials at levels A and B and 3 trials at levels C and D fall outside it. Single-level panels include 9 (NV), 10 (sSPVI) and 17 (SPVI) trials, and pooled panels 18, 20 and 34.

## Notes

### Clinical Trial

NCT05028712

### Author Declarations

The Johns Hopkins Medicine Institutional Review Board gave ethical approval for this work.

